# Assessing Relative Coronavirus Disease 2019 Mortality: A Swiss Population-based Study

**DOI:** 10.1101/2020.06.10.20127670

**Authors:** Torsten Hothorn, Matthias Bopp, Huldrych F. Günthard, Olivia Keiser, Maroussia Roelens, Caroline E. Weibull, Michael J. Crowther

## Abstract

**Objective:** Severity of the coronavirus disease 2019 (covid-19) has been previously reported in terms of absolute mortality in SARS-CoV-2 positive cohorts. An assessment of mortality relative to mortality in the general population is presented.

**Design:** Retrospective population-based study.

**Setting:** Individual information on symptomatic confirmed SARS-CoV-2 patients and subsequent deaths from any cause were compared to the all-cause mortality in the Swiss population of 2018. Starting February 23, 2020, mortality in covid-19 patients was monitored for 80 days and compared to the population mortality observed in the same time-of-year starting February 23, 2018.

**Participants:** 5 169 071 inhabitants of Switzerland aged 35 to 95 without covid-19 (general population in spring 2018) and 20 769 persons tested positively for covid-19 during the first wave in spring 2020.

**Measurements:** Sex- and age-specific mortality rates were estimated using Cox proportional hazards models. Absolute probabilities of death were predicted and risk was assessed in terms of relative mortality by taking the ratio between the sex- and age-specific absolute mortality in covid-19 patients and the corresponding mortality in the 2018 general population.

**Results:** Absolute mortalities increased with age and were higher for males compared to females, both in the general population and in positively tested persons. A confirmed SARS-CoV-2 infection substantially increased the probability of death across all patient groups at least nine-fold. The highest relative risks were observed among males and younger patients. Male covid-19 patients exceeded the population hazard for males (hazard ratio 1.23, 1.03 to 1.47). An additional year of age increased the population hazard in covid-19 patients only marginally (hazard ratio 1.01, 1.00 to 1.02).

**Conclusions:** Health care professionals, decision makers, and societies are provided with an additional population-adjusted assessment of covid-19 mortality risk. In combination with absolute measures of risk, the relative risks presented here help to develop a more comprehensive understanding of the actual impact of covid-19.

**Strengths and limitations of this study:**

- An assessment of covid-19 risk in terms of relative mortality rates comparing a cohort of symptomatic and confirmed covid-19 patients to the general population is provided.
- Relative mortality for quantifying sex- and age-specific covid-19 risk is unaffected by seasonal effects, cause of death (covid-19 related or not), impact of public health interventions, or testing coverage.
- The study excluded patients younger than 35 years and older than 95 years, as well as patients tested posthumously or after having been admitted to a hospital for reasons other than covid-19.
- Information about the distribution of relevant comorbidities was not available on population level and the associated risk was not quantified.
- Differential covid-19 risk for relevant patient subpopulations, such as pregnant women, low-income residents, ethnic or cultural minorities, was not assessed.

## Introduction

Early reports from China and Italy^1–7^ on disease severity of covid-19 caused unprecedented public health interventions around the world, ranging from social distancing measures or school and university closings to complete lockdowns of societies. The absolute mortality in patients diagnosed with covid-19, i.e., the case-fatality rate, has been the main entity used for communicating risks associated with the disease^2–6^. Risk factors for mortality risk, most prominently higher age, being male, and preexisting medical conditions, have become publically known^1,3,4^.

On the current occasion of liberalisation of the most stringent public health interventions in many countries, an assessment of the actual impact of the covid-19 pandemic is called-for. Health care professionals, politicians, and societies at large currently engage in a discussion about the appropriateness of the mitigation measures taken, and the first scientific contribution on the matter has arisen^8^.

The probability of death estimated from hundreds of thousands of covid-19 patients are constantly reported from many countries^2–4,9^. These numbers can, however, be hard to compare, due to differences in testing regimes, varying ascertainment of mortality, different age structures of societies, or different health-care and public health systems^10^. As an alternative risk difference of disease impact, excess numbers of deaths has been reported for some populations, i.e., the number of observed all-cause deaths during the time of the covid-19 pandemic (end of February to mid-May 2020 in most European countries) minus the expected number of deaths in the given population. Reports on the number of excess deaths observed since the onset of the pandemic are available from Portugal^11^, Spain^12^, northern Italy^13,14^, various other European countries^15^, and the United States^16^. However, cross-country comparisons are again difficult, because the success or failure of public health interventions and possible overruns of hospital capacities will be reflected in the presence and magnitude of excess mortality^14^.

The emergence of mature data on the course of the covid-19 pandemic from many countries allows its actual impact on societies and health-care systems to be discussed in light of the shortterm relative mortality. This relative risk compares the absolute all-cause mortality observed in patients diagnosed with covid-19 during the spring 2020 outbreak with the absolute all-cause mortality in the uninfected population of earlier years during the same calendar time of year. The population mortality varies between females and males, and over attained age, with males and older individuals experiencing a higher short-term risk of dying. How much of the increased mortality reported for male and older covid-19 patients^1,4,6,7^ can be attributed to the increase in population mortality risk in general is an important question awaiting an answer. Furthermore, sex- and age-specific relative covid-19 mortality allows a stratified assessment of risk-increase caused by a SARS-CoV-2 infection^17^.

We report an assessment of age-adjusted relative covid-19 mortalities for females and males based on an analysis of individual population level and covid-19 death records from Switzerland covering the time between 2020-02-24 and 2020-05-14.

## Methods

### Study design and data sources

For this population-based study, Swiss general population data from 2014–2018, including individual death records, were obtained from the Swiss Federal Statistical Office (Bundesamt für Statistik). In addition, covid-19 surveillance reports from the Swiss Federal Office of Public Health (Bundesamt für Gesundheit) on an unselected group of individuals tested positively for SARS-CoV-2 during the first wave between 2020-02-24 and 2020-05-14 were available, also including individual dates of tests and occurred deaths.

Official SARS-CoV-2 testing in Switzerland was performed by polymerase chain reaction (PCR) only, based on lower and upper respiratory tract samples from symptomatic persons. Individuals experiencing the following symptoms were eligible for testing^18^: cough, sore throat, muscle pain, dyspnea (with or without fever), and acute anosmia or ageusia. Testing of asymptomatic persons was only recommended to control local outbreaks in hospitals or nursing homes. A number of hospitals started to test all patients admitted to the hospital at different time points regardless of symptoms. Information of whether or not a person experienced symptoms during an infection was not available for this analysis. All positive and negative SARS-CoV-2 test results were directly reported to the Federal Office of Public Health, patients were followed-up subsequently, and all positive cases were included in this analyis.

### Study population

The study population consists of two cohorts. The first cohort consists of persons with a SARS-CoV-2 positive PCR test between 35 and 95 years of age at time of testing. We excluded younger covid-19 patients because no deaths were observed in this group. Individuals tested post-mortem or after hospitalisation for other reasons were not included as the aim was to study relative mortality in a cohort of newly infected people who did not have a short-term increased mortality risk. Very old persons (older than 95) years were also excluded for this reason. We refer to this cohort as the “Swiss covid-19” cohort in the sequel. The second cohort was defined as all inhabitants of Switzerland alive on February 23, 2018 aged 35 to 95 years.

Table 1 describes the selection process defining the study population. For an additional sensitivity analysis, the Swiss 2014 to 2017 study populations were defined analogously.

**Table 1:**
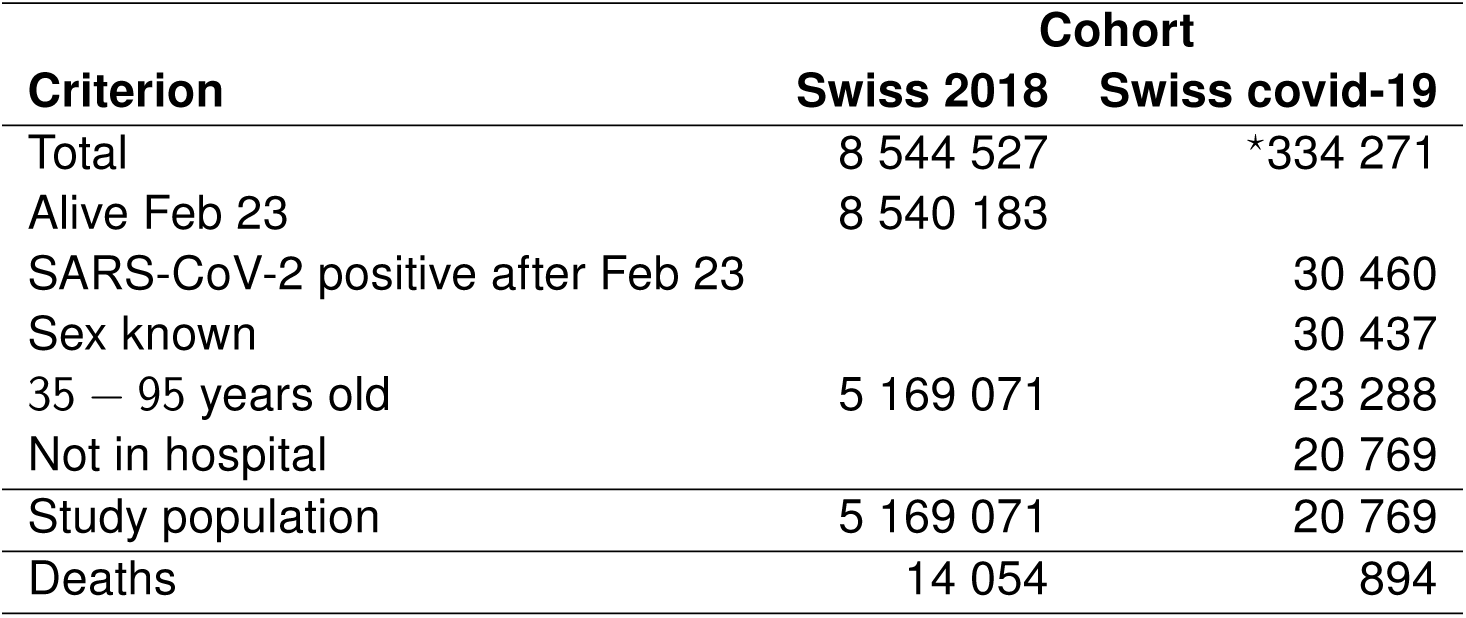
2018 study population. Swiss 2018 population cohort (as of January 1st, 2018) and Swiss covid-19 cohort (SARS-CoV-2 positive cases in a total of 334’271 tests performed in Switzerland between 2020-02-24 and 2020-05-14). The table contains the number of persons meeting the inclusion criteria. “Study population” refers to the number of observations in the two cohorts analysed. ^*^The total number of tests includes multiple counts of persons tested more than once.

| **Criterion** | **Cohort** |  |
| --- | --- | --- |
|  | **Swiss 2018** | **Swiss covid-19** |
| Total | 8 544 527 | *334 271 |
| Alive Feb 23 | 8 540 183 |  |
| SARS-CoV-2 positive after Feb 23 |  | 30 460 |
| Sex known |  | 30 437 |
| 35 – 95 years old | 5 169 071 | 23 288 |
| Not in hospital |  | 20 769 |
| Study population | 5 169 071 | 20 769 |
| Deaths | 14 054 | 894 |

### Statistical analysis

Exploratory analyses were performed comparing the sex- and age distributions between the Swiss population cohort and the Swiss covid-19 cohort. Absolute numbers and ratios were computed for females and males. In addition to mean ages (Table 2), nonparametric density estimates of age stratified by sex were computed and compared between the two cohorts (Figure 2).

**Table 2:**
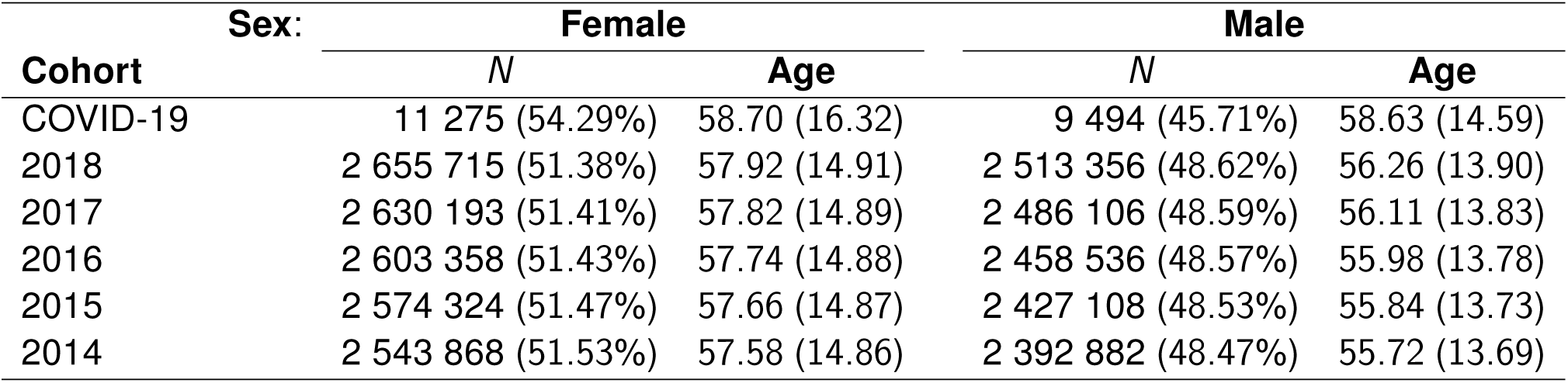
Study populations. Number of observations (*N*) and age (mean and standard deviation) for females and males 35 to 95 years old and alive on February 23 of the respective year. The first row corresponds to the Swiss covid-19 cohort of 2020.

**Figure 1:**
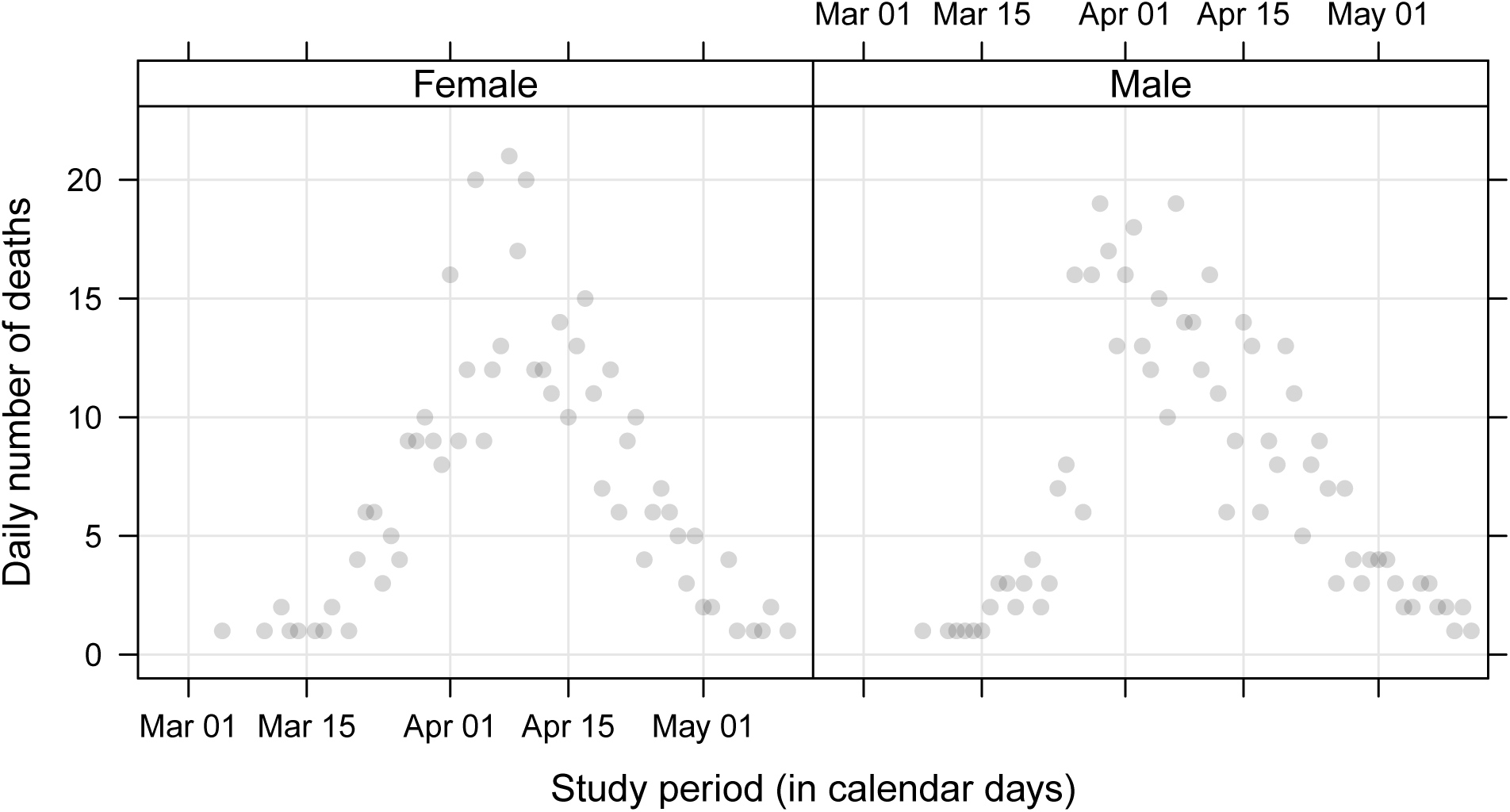
Swiss covid-19 cohort. Number of deaths from all causes reported between 2020-02-24 and 2020-05-14 in covid-19 patients (35 and 95 years old, not admitted to a hospital prior to testing, excluding post mortem tests).

**Figure 2:**
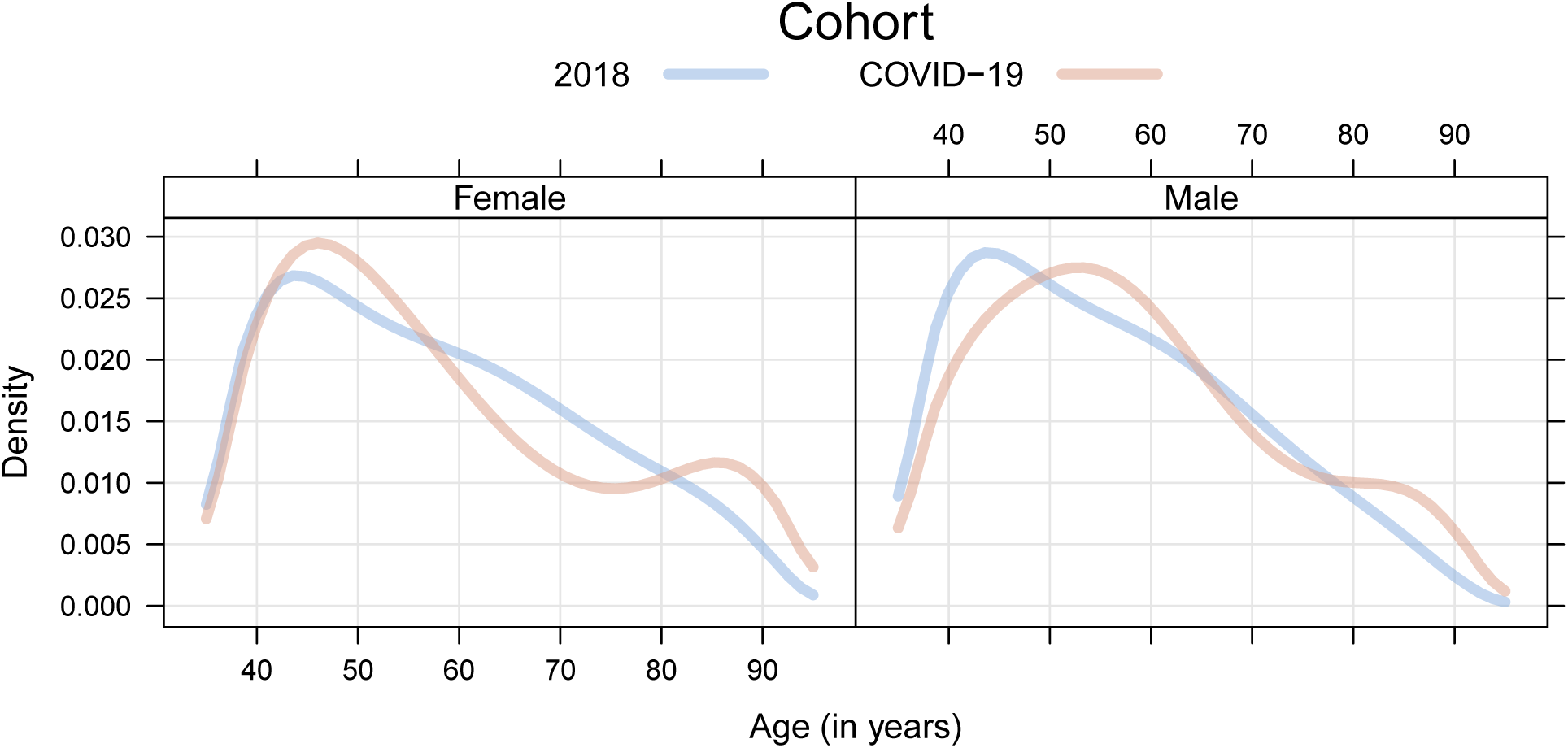
Swiss 2018 population and covid-19 cohorts. Comparison of age densities between the Swiss 2018 population and covid-19 cohorts, separately for females and males.

Mortality data was analysed using survival analysis. For the Swiss population cohort, follow-up started on February 23, 2018, and ended at date of death or on date of administrative censoring (May 14, 2018). The underlying time scale for all analyses was time since February 23, measured in days. The Swiss covid-19 cohort was handled in two different ways. Throughout, follow-up started on the day of the positive test and ended at time of death or on date of administrative censoring (May 14, 2020). When estimating relative rates (contrasting to the general population), the underlying time scale was time since February 23 (measured in days), using a delayed entry approach. For predicting the probability of death, number of days since positive test was used as the underlying time scale.

Sex- and age-specific hazard ratios (HRs) with 95% confidence intervals (CIs) were estimated using a stratified Cox proportional hazards model, allowing for separate baseline rates in the Swiss covid-19 and Swiss 2018 population cohorts, with 65-year old females as reference. Sex and age were modelled with main effects only for the Swiss 2018 cohort, whereas for the Swiss covid-19 cohort, interaction effects between SARS-CoV-2 status and sex and age were included to capture the additional mortality effects among the patients. *P*-values and 95% confidence intervals for hazard ratios were adjusted for multiplicity (see Online Supplement).

To quantify the impact on mortality associated with a SARS-CoV-2 diagnosis, the 60-day probability of death was predicted from the fitted model estimates, which captures deaths occurring within 60 days of February 23 or positive test, for the Swiss 2018 population cohort and Swiss covid-19 cohort, respectively. Using these probabilities, the sex-specific relative mortality (and associated 95% confidence bands) was calculated by taking the ratio between the two, along an age gradient between 35 and 95 years. The relative mortality incorporated uncertainty in the Swiss covid-19 cohort only^19^.

The assumption of proportional hazards was assessed by fitting models allowing for time-varying effects. Potential deviations from the linear age effect were assessed in a Cox model allowing nonlinear effects of age. The main-effects only model was compared to a model including sex *×* age interactions. As a sensitivity analysis, all models were re-fitted using the Swiss 2014 to 2017 general populations as reference. Further details on the statistical analyses performed can be found in the Online Supplement.

All analyses were performed in the R system for statistical computing^20^ (version 4.0.2) with the **survival**^21^ and **mlt**^22,23^ add-on packages and independently replicated in Stata^24^ (version 16), using the **stpm2**^25,26^ and **merlin**^27^ commands.

## Results

The daily number of deaths observed during the 80-day study period (2020-02-24 to 2020-05-14) in the Swiss covid-19 cohort increased rapidly from mid-March and peaked during the first days of April, in both males and females (Figure 1). The numbers reduced to less than ten reported deaths per day during the last week of the study period.

Characteristics of the Swiss covid-19 cohort, the Swiss 2018 population cohort, and the earlier Swiss population cohorts (2014-2017) are presented in Table 2. The ratio of females to males indicate that patients in the Swiss covid-19 cohort were more likely female. The mean age was similar between the two cohorts, with SARS-CoV-2 positive patients being between one (females) and up to three years (males) older than individuals in the Swiss population cohort. The similarity was further observed when comparing the whole age distribution among females and males, between cohorts (Figure 2). However, there was a slight over-representation of older people, between 85 and 95 years of age, in SARS-CoV-2 positive individuals.

The all-cause mortality rate among 65-year old males from the Swiss 2018 population cohort was 1.46 times that among females of the same age. The mortality rate further increased by a factor of 1.13 for each additional year of age (Table 3). In 2018, males experienced the same mortality as females aged 3.15 years older.

**Table 3:** Swiss 2018 population and covid-19 cohorts. Log-hazard ratios (log-HR) and hazard ratios (HR) expressing the risk of being male (“Male”) and each year of age (“Age – 65”) compared to the baseline hazard in 65 year old females. Main effects were fitted to both cohorts, interaction effects to the Swiss covid-19 cohort only. The interaction effects describe the additional risk on the log-HR or HR scale attributable to the infection. Estimates are given with standard errors (SEs) for log-HRs and 95% confidence intervals (CI) for HRs, the latter and *P*-values were adjusted for multiplicity.

| Effect | log-HR | SE×10 | *P*-value | HR | 95% CI |
| --- | --- | --- | --- | --- | --- |
| Female | 0 |  |  | 1 |  |
| Male | 0.38 | 0.17 | < 0.001 | 1.46 | 1.40–1.53 |
| Age 65 | 0 |  |  | 1 |  |
| Age – 65 | 0.12 | 0.01 | < 0.001 | 1.13 | 1.13–1.13 |
| covid-19 × Female | 0 |  |  | 1 |  |
| covid-19 × Male | 0.21 | 0.70 | 0.01 | 1.23 | 1.03–1.47 |
| covid-19 × Age 65 | 0 |  |  | 1 |  |
| covid-19 × Age – 65 | 0.01 | 0.03 | 0.10 | 1.01 | 1.00–1.02 |

The additional increment in the mortality rate for individuals in the Swiss covid-19 cohort were smaller than the sex- and age-effects observed in the Swiss 2018 population cohort (difference in log-HRs between population and patients 0.17, 95% CI: 0.00 − 0.34, for the sex effect and 0.11, 95% CI: 0.11 − 0.12, for the age effect). Being a male covid-19 patient was associated with a HR of 1.23, relative to males in the general population (Table 3). The hazard ratio comparing the hazard of males to females in the Swiss covid-19 cohort was 1.46 *×* 1.23 = 1.80 (HR, 95% CI: 1.58 − 2.06). On the log-hazard ratio scale, the sex effect for male covid-19 patients is 0.38 + 0.21 = 0.59 and thus roughly two-thirds of the increased risk in a direct comparison of male to female covid-19 patients can be attributed to the higher mortality of males in the general population. The confidence interval for the covid-19 *×* Male term (HR 1.23, 95% CI: 1.03 − 1.47) reflects a substantial uncertainty regarding the prognostic relevance of being male with respect to covid-19 mortality.

Each additional year of age increased the covid-19 mortality rate by a factor of 1.13 *×* 1.01 = 1.14 (HR, 95% CI: 1.13−1.14). Formulated alternatively, the age-related hazard doubled every 5.75 life years in the Swiss 2018 population cohort and every 5.42 life years in the Swiss covid-19 cohort. This additional effect of age in covid-19 patients was rather small (HR, 95% CI: 1.00 − 1.02) and not significant at the 5% level. Hazards of male patients were comparable to hazards of female patients 4.60 years older.

The 60-day sex- and age-specific probability of death (absolute mortality) was considerably larger in the covid-19 cohort (Figure 3, top panel and Table 4). The absolute number of expected deaths per 100000 persons within 60 days ranged from 3 and 4 (35 year old females and males) to 3658 and 5303 (95 year old females and males) in the 2018 population cohort. In covid-19 patients, these numbers increased to 43 and 82 expected deaths in 35 year old females and males and to 38127 and 60177 expected deaths in 95 year old females and males.

**Table 4:** Swiss 2018 population and covid-19 cohorts. Absolute and relative mortalities. Estimated number of deaths per 100 000 females or males with corresponding age. Population mortality in the Swiss 2018 cohort (Absolute 2018), mortality in the Swiss covid-19 cohort (Absolute SARS-CoV-2), and the relative SARS-CoV-2 mortality (RM, the ratio of the second to the first column). Confidence intervals were obtained from 95% confidence bands given in Figure 3.

| Sex | Age | Mortality |  |  |
| --- | --- | --- | --- | --- |
|  |  | Absolute 2018 | Absolute SARS-CoV-2 | Relative (RM) |
| Female | 35 | 3 (2–3) | 43 (28–65) | 16 (10–24) |
|  | 40 | 5 (4–5) | 77 (52–113) | 16 (11–23) |
|  | 45 | 9 (8–10) | 138 (97–196) | 15 (11–22) |
|  | 50 | 16 (15–18) | 247 (180–339) | 15 (11–21) |
|  | 55 | 30 (28–32) | 443 (334–587) | 15 (11–20) |
|  | 60 | 55 (52–58) | 794 (619–1 018) | 14 (11–19) |
|  | 65 | 100 (95–105) | 1 421 (1 142–1 767) | 14 (11–18) |
|  | 70 | 183 (176–190) | 2 538 (2 095–3 073) | 14 (11–17) |
|  | 75 | 334 (322–346) | 4 511 (3 803–5 348) | 14 (11–16) |
|  | 80 | 609 (589–630) | 7 955 (6 798–9 299) | 13 (11–15) |
|  | 85 | 1 110 (1 073–1 148) | 13 831 (11 889–16 062) | 12 (11–14) |
|  | 90 | 2 019 (1 945–2 095) | 23 458 (20 166–27 191) | 12 (10–13) |
|  | 95 | 3 658 (3 505–3 817) | 38 127 (32 784–44 021) | 10 (9–12) |
| Male | 35 | 4 (4–4) | 82 (55–121) | 21 (14–31) |
|  | 40 | 7 (7–8) | 147 (103–210) | 20 (14–29) |
|  | 45 | 13 (12–14) | 264 (192–363) | 20 (15–28) |
|  | 50 | 24 (22–26) | 473 (356–627) | 20 (15–26) |
|  | 55 | 44 (41–46) | 848 (661–1 086) | 19 (15–25) |
|  | 60 | 80 (76–84) | 1 517 (1 222–1 883) | 19 (15–24) |
|  | 65 | 146 (140–153) | 2 708 (2 245–3 264) | 19 (15–22) |
|  | 70 | 267 (257–277) | 4 810 (4 084–5 662) | 18 (15–21) |
|  | 75 | 488 (471–504) | 8 472 (7 314–9 804) | 17 (15–20) |
|  | 80 | 889 (860–919) | 14 698 (12 795–16 857) | 17 (14–19) |
|  | 85 | 1 619 (1 562–1 678) | 24 836 (21 655–28 394) | 15 (13–18) |
|  | 90 | 2 938 (2 822–3 059) | 40 112 (35 000–45 676) | 14 (12–16) |
|  | 95 | 5 303 (5 063–5 555) | 60 177 (52 995–67 468) | 11 (10–13) |

**Figure 3:**
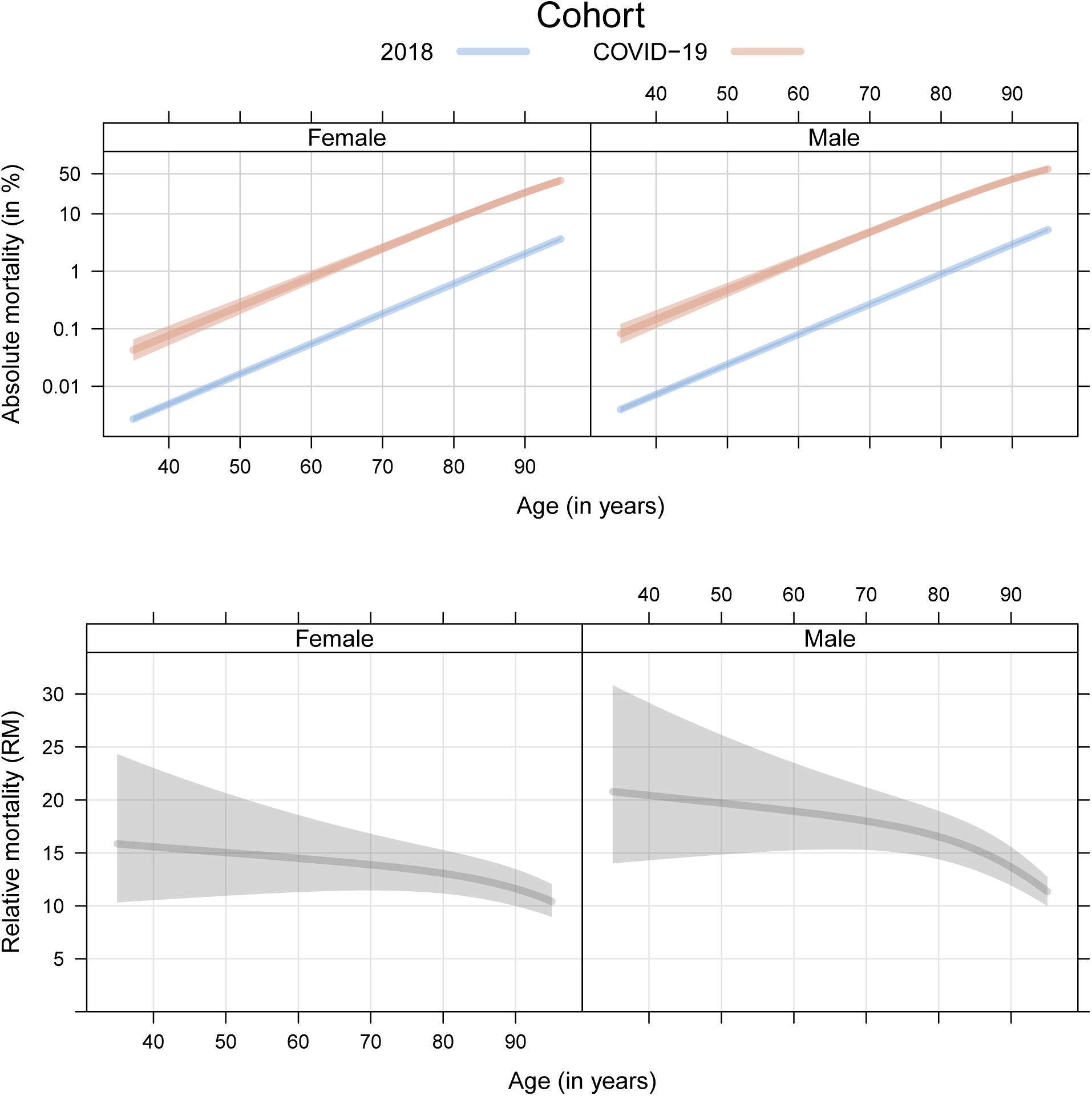
Swiss 2018 population and covid-19 cohorts. Comparison of absolute mortality (the probability of dying from any cause after 60 days, plotted on logarithmic scale) between the Swiss 2018 population and covid-19 cohorts, separately for females and males of different ages (top). Comparison of relative mortality (RM, the ratio of absolute mortalities in the covid-19 versus the 2018 cohort) between the two cohorts, for females and male of different ages (bottom). All estimates are plotted with 95% confidence bands.

The short-term death probability for 42-year old females diagnosed with covid-19 was about 0.1%, comparable with 65-year old females in the 2018 population cohort. Similarly, 55-year old infected males were associated with a probability of death around 1%, equivalent to the probability of 81-year old not infected males in the general population. The probability of death in infected individuals was above 10% for females older than 82 and males older than 76 years.

The probabilities of death implied a nearly log-linear function of age, for both females and males, and in both cohorts of the study population. Hence, it follows that the relative risk comparing the mortality in young patients to the mortality in patients aged 20 years older, for example, would be constant regardless of the age of younger group of patients. The slope of the probabilities of death as a function of age was about the same in the Swiss covid-19 cohort due to the models’ negligible age effect attributable to the infection (Table 3, covid-19 *×*Age − 65).

The sex- and age-specific relative mortality (Figure 3 bottom panel and Table 4) was largest in young male patients and declined with increasing age for both females and males. A relative mortality of 15 (in 50 year old females) is interpreted as 50 year old infected females being 15 times more likely to die within 60 days after the positive test than 50 year old uninfected females were likely to die within 60 days following February 23, 2018. However, the 95% confidence bands demonstrated substantial uncertainty in the estimates of this relative risk; the lower confidence band was as large as 9 for 95-year old females and 10 for 95-year old males. The data were consistent with an at least 11-fold risk increase in younger females (40 to 85 years old) and with an at least 15-fold risk increase in younger males (45 to 75 years old).

Sensitivity analyses (see Online Supplement) demonstrated that results reported for the 2018 study population were very close to the results for the 2014 to 2017 study populations. Model diagnostics did not reveal relevant deviations from proportional hazards nor the main effects model, but indications of a nonlinear age effect in the Swiss covid-19 cohort were found. The nonlinear model suggested a continuously and monotonically increasing hazard of age lacking any clear cut-off points for binary risk stratification.

## Discussion

The at least nine fold increase in probability of death found in female and male, young and old, symptomatic covid-19 patients from Switzerland in comparison to the Swiss population of 2018 provides novel sex- and age-specific information on the severity of this pandemic. Short-term covid-19 mortality has never previously been reported as a relative risk in direct relation to the short-term mortality in the general population. The comparison of absolute mortalities and case-fatality rates between risk groups of covid-19 patients without any population adjustment are likely to over-estimate the increased mortality in males and older people. As presented here, two-thirds of the risk increase observed in male patients could be attributed to the generally increased population mortality.

The population-based setting with matched calendar time allows for estimation of covid-19 related mortality in an unselected cohort of symptomatic and diagnosed patients, without the need to differentiate between deaths caused or not caused by covid-19 in terms of death certificate information^28^. Seasonal effects were implicitly accounted for by comparing cohorts over the same time-of-year. Unlike other population studies estimating excess mortality, the probability of death in these cohorts could not be confounded by ongoing public health interventions or testing coverage^28^. Using data from Switzerland was especially useful for this type of analysis. The borders to northern Italy and Austria caused covid-19 outbreaks early in the pandemic, broad and uniform symptom-based testing overseen by federal authorities was implemented quickly, and all test results were reported. A symptom-based testing regime was carried out, with testing of asymptomatic persons only recommended to control local outbreaks in hospitals or nursing homes. In contrast to reports from northern Italy^14^, the number of covid-19 patients in need for hospitalisation never exceeded health-care capacities, and every patient received the best possible treatment under the circumstances.

Absolute mortalities in the Swiss covid-19 cohort were smaller than those reported for Italian and Chinese covid-19 patients^3–5^ between 40 and 80 years old. The numbers are expected to be higher than in Germany, where the overall case fatality rate was only 1.2% and more tests were performed in younger patients with mild symptoms^29^. Due to differences in testing protocol, substantially higher or lower relative mortality may therefore be expected in other countries. The Swiss covid-19 cohort excluded persons with known increased mortality risks (those tested posthumously or while being hospitalised), as well as very old and thus a priori frail persons. The figures presented here can nevertheless inform models developed for computing prognoses on the number of expected deaths in real or hypothetical populations, because relative mortality is not affected by public health interventions which lead to a reduced or even nonexistent excess mortality in many European countries. For the UK, prognoses assumed a 1-year relative mortality risk not larger than two, uniformly for females and males of all ages^30^. A short-term 60-day relative mortality larger than nine, as found here, suggests that the actual risk might be larger than assumed based on prognostic models or reported elsewhere^31^. However, the more general question of the true covid-19 relative mortality will be lower than that reported in this study, as the inclusion criteria for the covid-19 cohort was defined on being symptomatic and testing positive for SARS-CoV-2. The true proportion of asymptomatic cases and the actual prevalence are ongoing questions^32^.

The sex- and age effects on all-cause mortality attributable to covid-19 are, however, still relevant and males and older persons were associated with higher risk. Males had a 23% higher mortality rate than females, however, the uncertainty around this hazard ratio was large and the effect was absent in some populations of earlier years and in a model with sex *×* age interactions (see Online Supplement). The relative mortality (ratio of the probabilities of death) was approximately between ten and twenty across all age groups. This population-adjusted comparison of the risk between young and old patients suggest a less drastic relative impact of age on mortality than previously reported^8^. Stratification into low and high risk groups using a cutoff at 65 years of age, as seen in several published reports and recently advocated for^33^, seems hard to justify based on the continuously increasing age-specific absolute mortalities and almost constant relative mortalities reported here. Relative mortality can also help to explain an increased mortality in younger patients reported from countries with different age structure: In Chile, 8% of those who died after an SARS-CoV-2 infection were less than 50 years old^34^, a number much higher than observed in Europe. Potential deviations from these sex- and age-specific covid-19 risks in relevant patient subgroups, such as patients with preexisting conditions^1^, pregnant women^35^, and racial, ethic, or cultural minorities^36^, were not modelled because of lack of information on related variables.

In summary, the results suggest that covid-19 risk assessment in terms of case-fatality rates and excess mortalities should be complemented by population-adjusted relative mortalities such that a more complete picture can emerge, potentially leading to improvements for age-based risk stratification.

## Data Availability

All results can be reproduced from study cohorts, R and Stata computer code publically available from https://gitlab.switch.ch/torsten.hothorn/relative_covid-19_mortality

## Contributions

TH, MB, and MJC designed the study, preprocessed, and analysed the data. TH prepared an initial draft, all authors reviewed and improved early versions of the manuscript and approved the final version.

## Funding

This work was supported by Swiss National Science Foundation.

## Competing interests

TH has been paid for consulting, lectures or presentations from Novartis, Roche, and PricewaterhouseCoopers. HFG has received unrestricted research grants from Gilead Sciences and Roche; fees for data and safety monitoring board membership from Merck; consulting/advisory board membership fees from Gilead Sciences, Merck and ViiV Healthcare; and grants from SystemsX, and the National Institutes of Health. MJC has been paid for consulting, lectures or presentations from Roche and Reinsurance Group of America UK Services Limited.

## Patient and public involvement

Patients and/or the public were not involved in the design, conduct, reporting or dissemination plans of this research.

## Patient consent for publication

Not required.

## IRB approval

Patient data was collected by the Swiss Federal Office of Public Health (Bundesamt für Gesundheit) under epidemic law.

## A. Online Supplement

### A.1. Statistical analysis

For the population cohort of a given year, all inhabitants of Switzerland alive and aged between 35 and 95 years old at February 23 enter the risk set. Inhabitants surviving 80 days were censored after 80 days, and those who died within 80 days after February 23 were recorded as an event. To be able to evaluate mortality rates and ratios on the same timescale, allowing for seasonal effects, for the covid-19 cohort, a delayed entry approach was taken, as patients do not become at risk of death until their date of positive test. All deaths in this cohort were treated as events. Individuals without a death record were administratively right-censored 80 days after February 23.

We therefore modelled time-to-death from any cause within 80 days after February 23 (the first positive SARS-CoV-2 test was recorded February 24, 2020; the database was closed on 2020-05-14). The mortality rate in the Swiss covid-19 cohort was compared to the mortality rate in the Swiss 2018 population cohort during the same time of year. Hazard ratios were estimated by a Cox proportional hazards model expressing the distribution of time-to-death after February 23 in days *t >* 0. We fitted a Cox model allowing separate baseline hazard functions, assuming

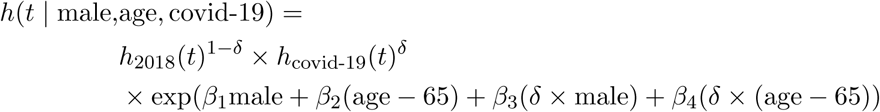

where *h*_2018_(*t*) is the baseline mortality rate at time *t*, for a 65 year old female in the Swiss 2018 population cohort, *h*_covid-19_(*t*) describes the mortality rate in the Swiss covid-19 cohort for (on the same calendar timescale to account for seasonal effects), and *δ* takes the value of 1 if the individual is in the covid-19 cohort, and 0 if the individual is in the 2018 cohort. The linear predictor is parameterised in such a way that we can quantify the effects of age and being male in the general population by the hazard ratios exp(*β*_1_) and exp(*β*_2_), respectively, and the *additional* increase/decrease in the hazard attributable to age and being male in the covid-19 cohort, described by the exp(*β*_3_) and exp(*β*_4_), respectively. The proportional hazards assumption was assessed by comparing the in-sample log-likelihood of the model described above to the in-sample log-likelihood of a Cox model with time-varying sex- and age-effects. The simpler model assuming proportional hazards maximised the log-likelihood at −5386·64 and the model not assuming proportional hazards at −5379·64, suggesting that mortalities were appropriately modelled under proportional hazards.

For the estimation and comparison of absolute risk, i.e. the probability of dying by time *t*, we model the mortality rate in the covid-19 cohort on the time since diagnosis timescale. Therefore, the model for the 2018 cohort remains the same, and we are interested in the probability of death 60 days after February 23, 2018. In the covid-19 cohort, we are interested in the probability of death 60 days after diagnosis. Two separate Cox models were fitted to the cohorts with the appropriate timescales. Relative mortality (RM), at time *t*, is then defined as,

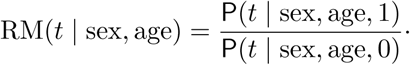

which is the ratio of the probability `P`(*t* | sex, age, 1) to die within *t* days after a positive SARS-CoV-2 test and the probability `P`(*t* | sex, age, 0) to die within *t* days after February 23, 2018 without an infection for female or male persons of a specific age. We used a time horizon of *t* = 60 days. A relative mortality RM(60 | female, 65) = *X* means that the probability to die within 60 after an SARS-CoV-2 infection is *X* times larger than the population probability to die within 60 days after February 23, 2018, for 65 year old females. Proportional hazards models for absolute risk were estimated with log-cumulative baseline hazard functions parameterised in terms of a Bernstein polynomial^22^, that is, using a parametric equivalent of the semiparametric Cox model.

In all models, the assumption of a linear age effect was assessed by comparing the linear Cox proportional hazards model to a Cox proportional model featuring a nonlinear additive effect of age on the log-hazard (Figure 1). For the 2018 population cohort, differences were marginal and justified the use of a linear age term. For the Swiss covid-19 cohort, the nonlinear model suggested a stronger age effect up to 80 years, which then levelled-off for older people. The discrepancy between the nonlinear and the linear Cox model in terms of the estimated probability of death at day 80 can be inferred from Figure 2. For the 2018 population cohort, the probability of death from both models were very similar, with some variation for people older than 90 years, which can be explained by the sharp increase of the corresponding log-hazard ratio (Figure 1). In the age range between 60 and 90 years, the two models were relatively similar (*±*25%), however, the nonlinear model suggested the probability of death was 50% reduced compared to probability of death from the linear model. This means that the relative mortalities assuming a linear effect of age on the log-hazard scale (Figure 3 and Table 4) could be up to two times smaller for people younger than 60 and up to two times larger for persons around 80 years old in a more complex model allowing a nonlinear effect of age. We also considered an interaction between sex and age, found significant effects yet hardly relevant effect sizes (HR, 0·99 for the 2018 population and 1·02 for the covid-19 cohort, see Table 1). It should be noted that the confidence interval for the hazard ratio of the covid-19 *×* Male effect is very wide and includes one. Thus, the interaction model suggests that all covid-19 patients suffer the same hazard increase, compared to the 2018 population.

### A.2. Computational details

Computations were performed in `R` version 4.0.2^20^. Cox proportional hazards models were fitted semiparametrically to estimate log-hazard ratios using the **survival**^21^ package. Absolute mortalities were obtained from Cox models, parametrically parameterised with flexible log-cumulative baseline hazard function, using package **mlt**^1^. Multiplicity adjusted *P* -values, confidence intervals, and confidence bands for Cox models were computed using the **multcomp** package^2^.

### A.3. Sensitivity analysis

The primary analyses were conducted in `R`; however, to ensure validity and robustness, all analyses were replicated independently in `Stata` by using a flexible parametric survival model^25^ using the **stpm2**^26^ and **merlin**^27^ commands, which can be considered a parametric equivalent to the Cox model but use restricted cubic splines to directly model the baseline (log cumulative) hazard function. We found complete agreement across software platforms, and modelling approaches.

**Supplementary Table 1:** Swiss 2018 population and covid-19 cohorts. Log-hazard ratios (log-HR) and hazard ratios (HR) expressing the risk of being male (“Male”) and each year of age (“Age−65”) compared to the baseline hazard in 65 year old females, based on a Cox proportional hazards model with sex *×* age interaction. Estimates are given with standard errors (SEs) for log-HRs and 95% confidence intervals (CI) for HRs, the latter and *P* -values were adjusted for multiplicity.

| Effect | log-HR | SE×10 | $P$-value | HR | 95% CI |
| --- | --- | --- | --- | --- | --- |
| Female | 0 |  |  | 1 |  |
| Male | 0.57 | 0.29 | < 0.001 | 1.76 | 1.64–1.90 |
| Age 65 | 0 |  |  | 1 |  |
| Age – 65 | 0.13 | 0.01 | < 0.001 | 1.14 | 1.13–1.14 |
| Male × Age – 65 | –0.01 | 0.02 | < 0.001 | 0.99 | 0.98–0.99 |
| covid-19 × Female | 0 | | | 1 | |
| covid-19 × Male | –0.12 | 1.39 | 0.87 | 0.89 | 0.62–1.26 |
| covid-19 × Age 65 | 0 | | | 1 | |
| covid-19 × Age – 65 | –0.00 | 0.05 | 0.92 | 1.00 | 0.98–1.01 |
| covid-19 × Male × Age – 65 | 0.02 | 0.07 | 0.01 | 1.02 | 1.00–1.04 |

**Supplementary Figure 1:**
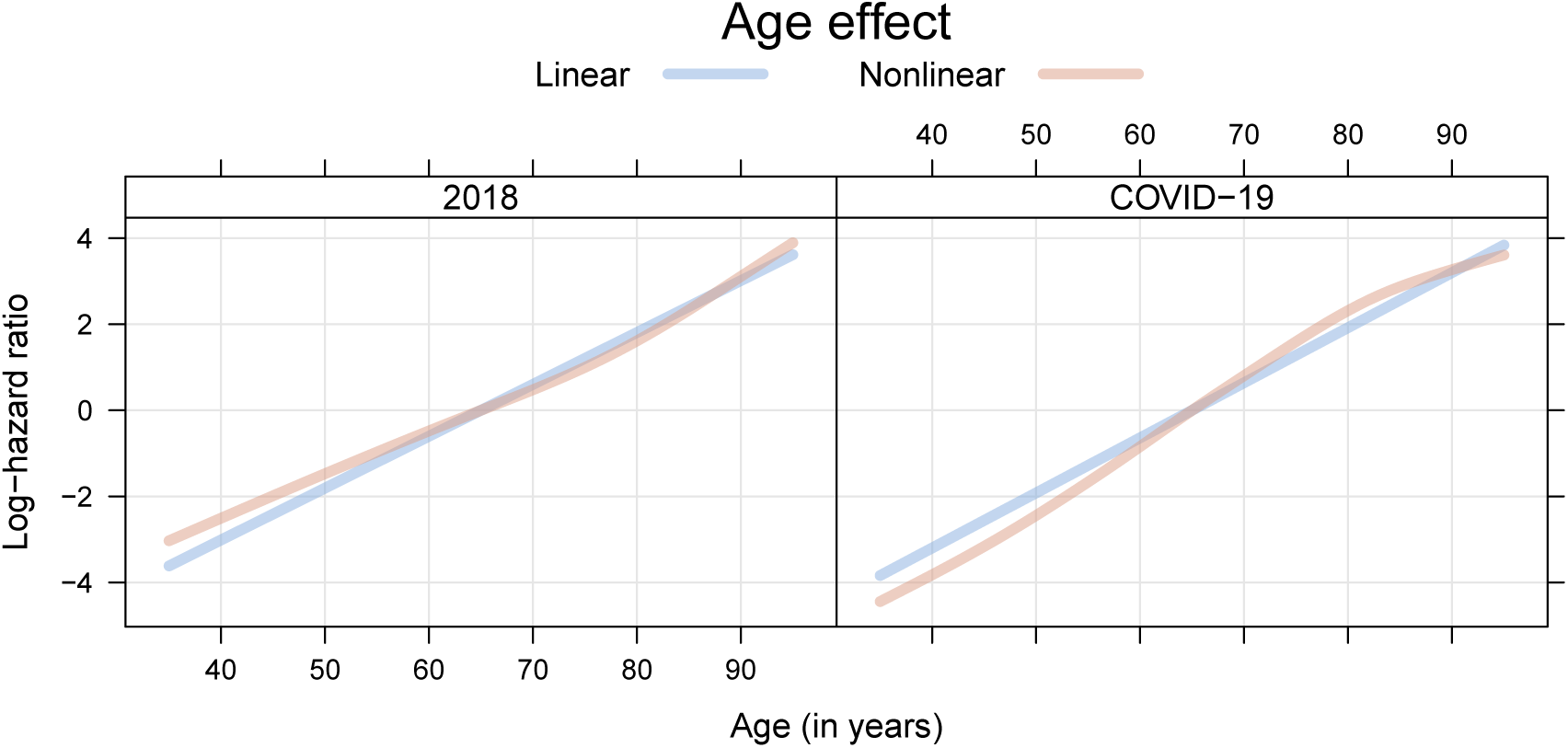
Swiss 2018 population and covid-19 cohorts. Evaluation of linearity assumption on age. A Cox proportional hazards model was refitted allowing a nonlinear additive impact of age on the log-hazard function. The age effects of the linear Cox model, for females in the two study cohorts, are compared on the log-hazard ratio scale.

**Supplementary Figure 2:**
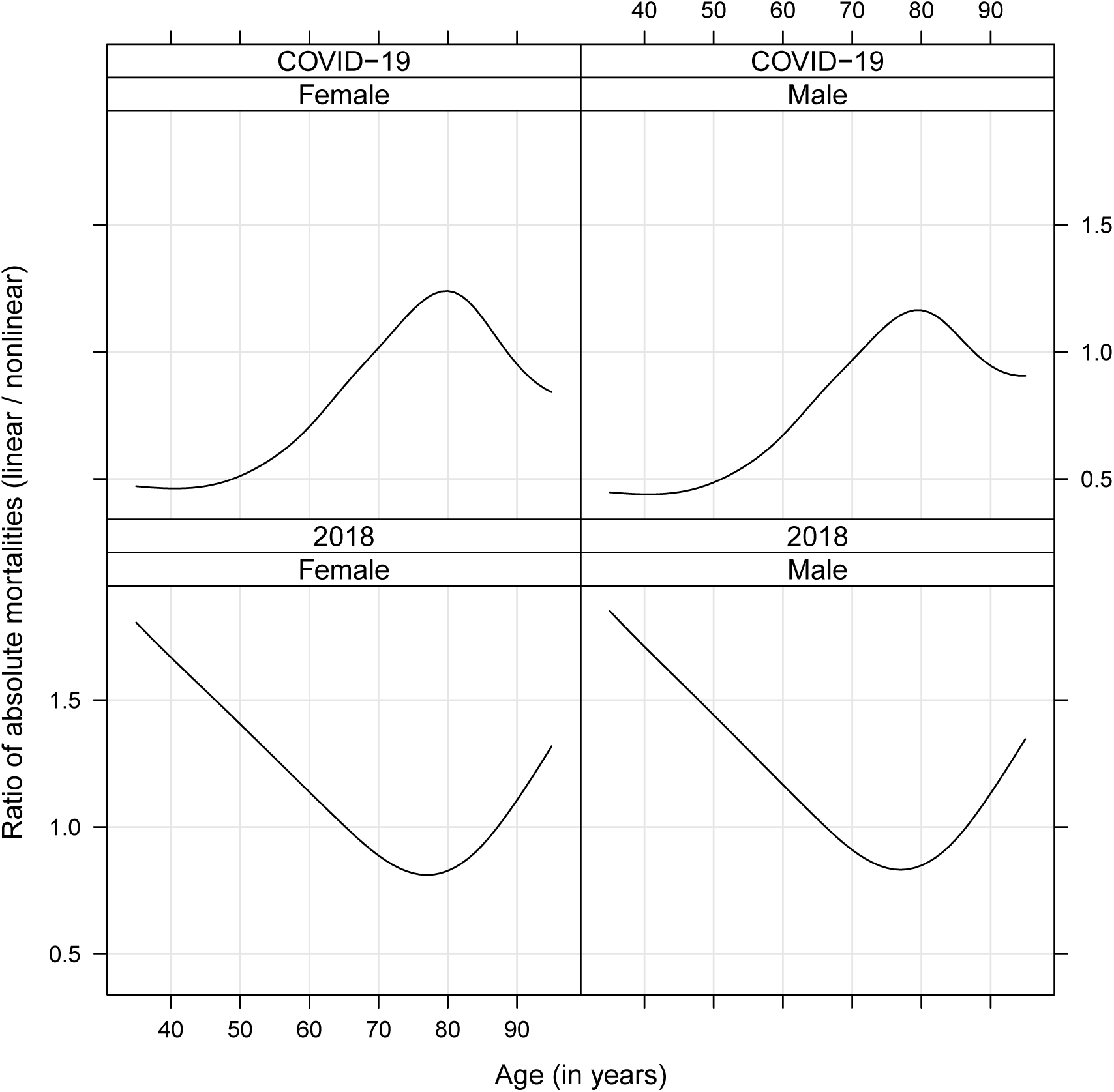
Swiss 2018 population and covid-19 cohorts. Comparison of linear and nonlinear Cox models. The ratio of absolute mortalities (nonlinear model vs. linear model) as a function of age, separately for females and males and the two study cohorts is displayed.

**Supplementary Figure 3:**
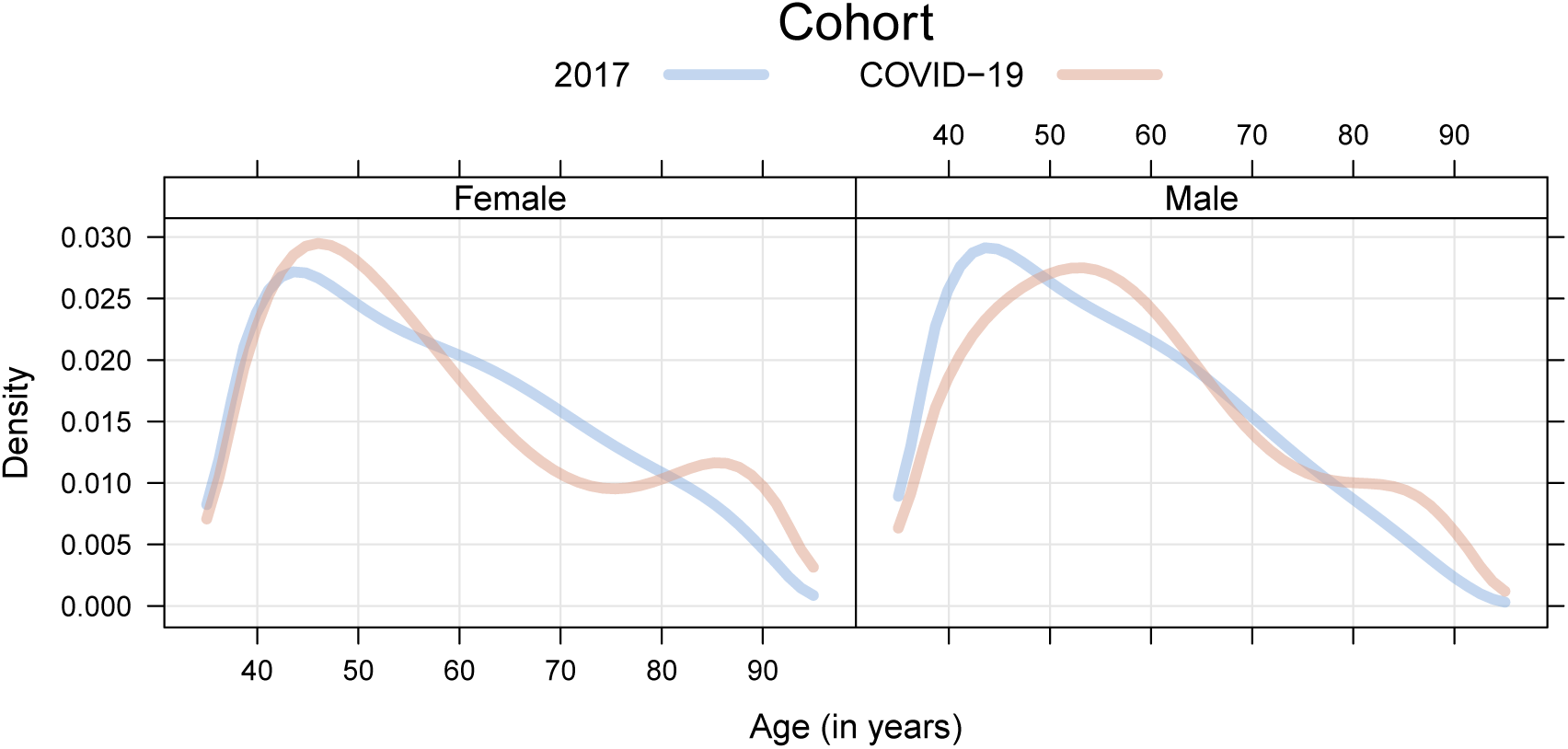
Swiss 2017 population and covid-19 cohorts. Comparison of age densities between the Swiss 2017 population and covid-19 cohorts, separately for females and males.

Furthermore, the sensitivity of the main findings reported was assessed by conducting the same analyses, but using the population data from 2014 to 2017, independently, as the comparative populations, instead of the 2018 cohort.

Age distributions and baseline cumulative hazards for these study populations are given in Figures 3, 5, 7, and 9. Absolute and relative mortalities can be obtained from Figures 4, 6, 8, and 10. Hazard ratios for all study populations are presented in Table 2. The deviations from the main results were marginal.

**Supplementary Table 2:**
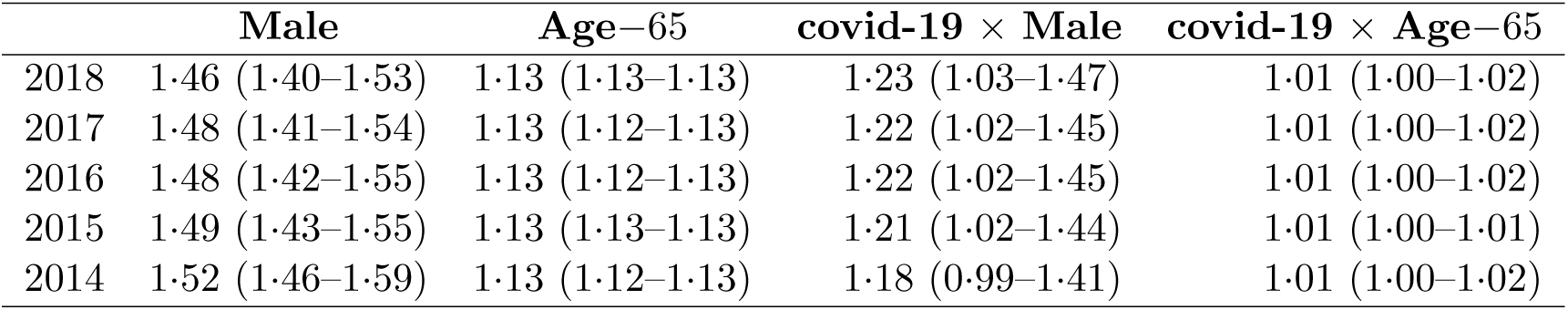
2014-2018 study populations. Hazard ratios expressing the risk of being male (“Male”) and each year of age (“Age − 65”) compared to 65 year old females. Each row corresponds to the study population consisting of the Swiss covid-19 cohort and the population cohort of the respective year.

**Supplementary Figure 4:**
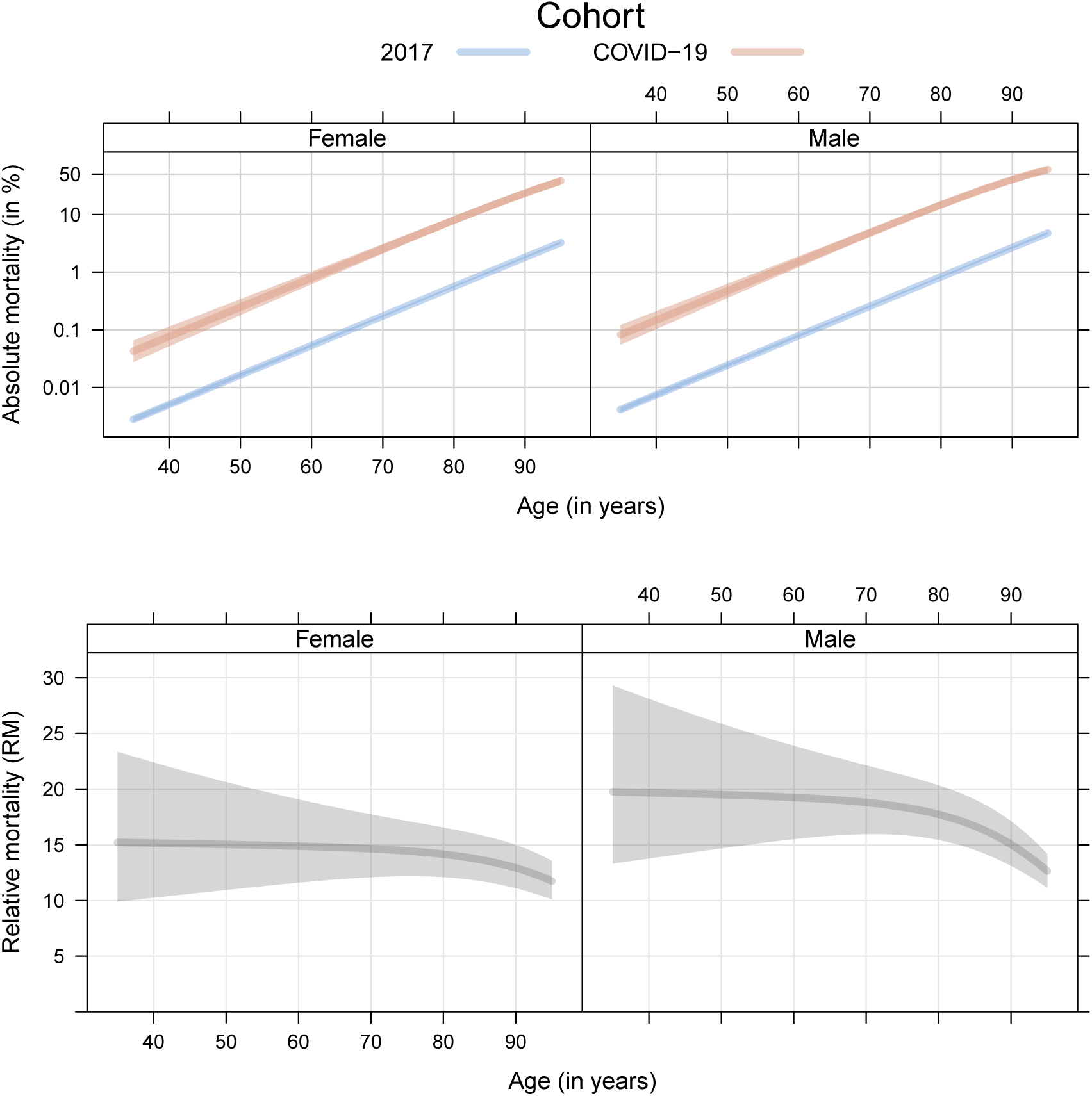
Swiss 2017 population and covid-19 cohorts. Comparison of absolute mortality (the probability of dying from any cause after 60 days, plotted on logarithmic scale) between the Swiss 2017 population and covid-19 cohorts, separately for females and males of different ages (top). Comparison of relative mortality (RM, the ratio of absolute mortalities in the covid-19 versus the 2017 cohort) between the two cohorts, for females and male of different ages (bottom). All estimates are plotted with 95% confidence bands.

**Supplementary Figure 5:**
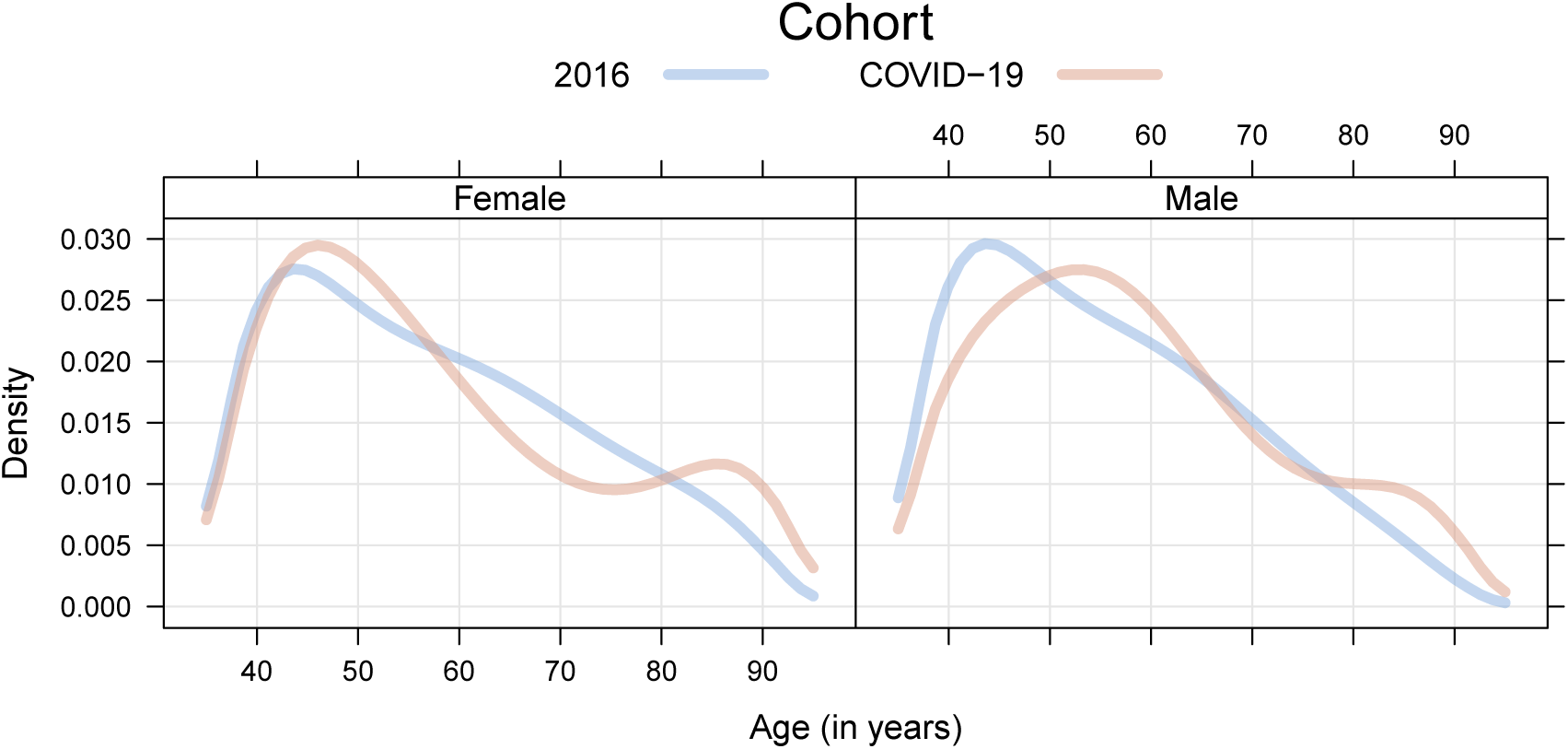
Swiss 2016 population and covid-19 cohorts. Comparison of age densities between the Swiss 2016 population and covid-19 cohorts, separately for females and males.

**Supplementary Figure 6:**
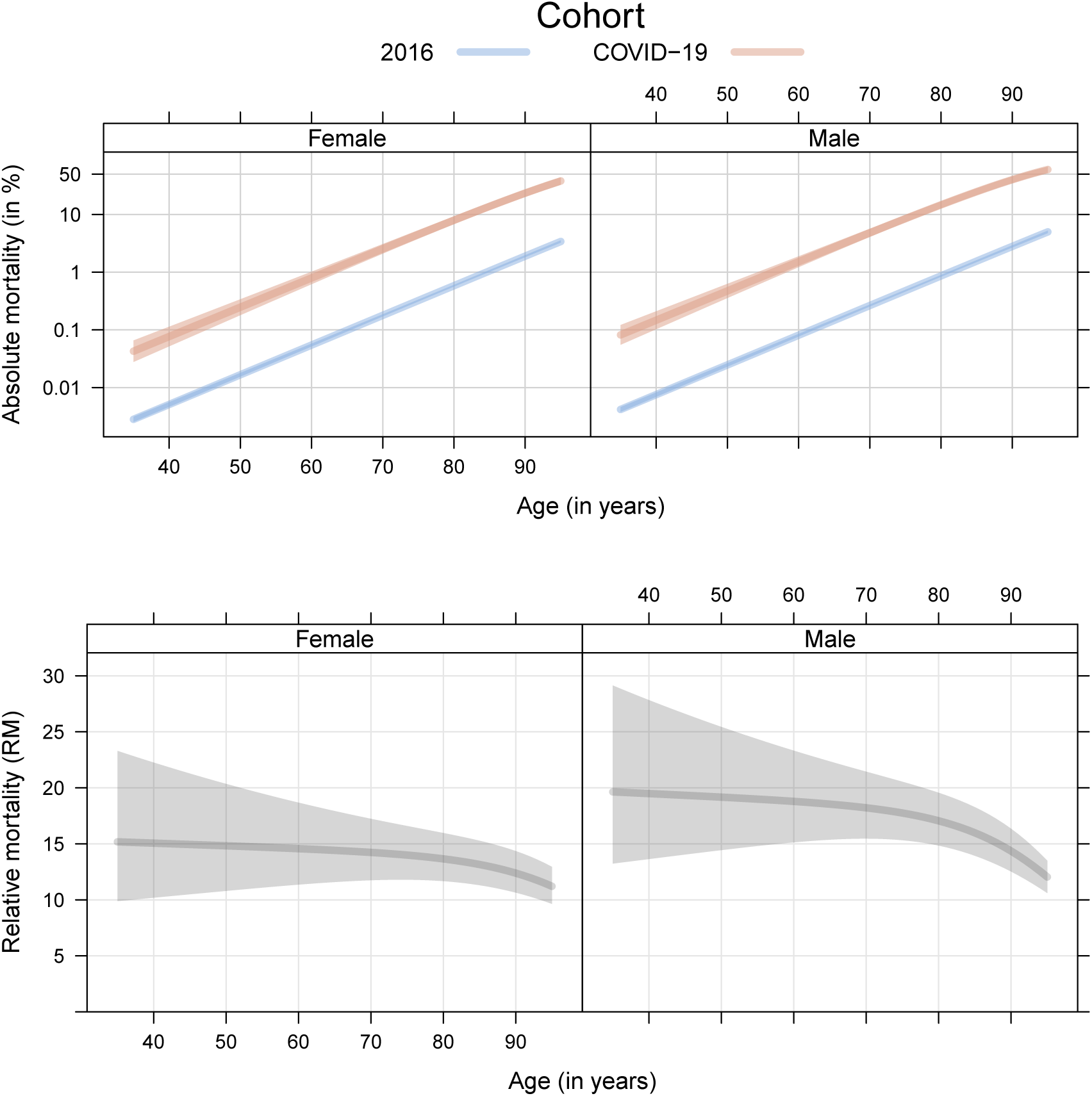
Swiss 2016 population and covid-19 cohorts. Comparison of absolute mortality (the probability of dying from any cause after 60 days, plotted on logarithmic scale) between the Swiss 2016 population and covid-19 cohorts, separately for females and males of different ages (top). Comparison of relative mortality (RM, the ratio of absolute mortalities in the covid-19 versus the 2016 cohort) between the two cohorts, for females and male of different ages (bottom). All estimates are plotted with 95% confidence bands.

**Supplementary Figure 7:**
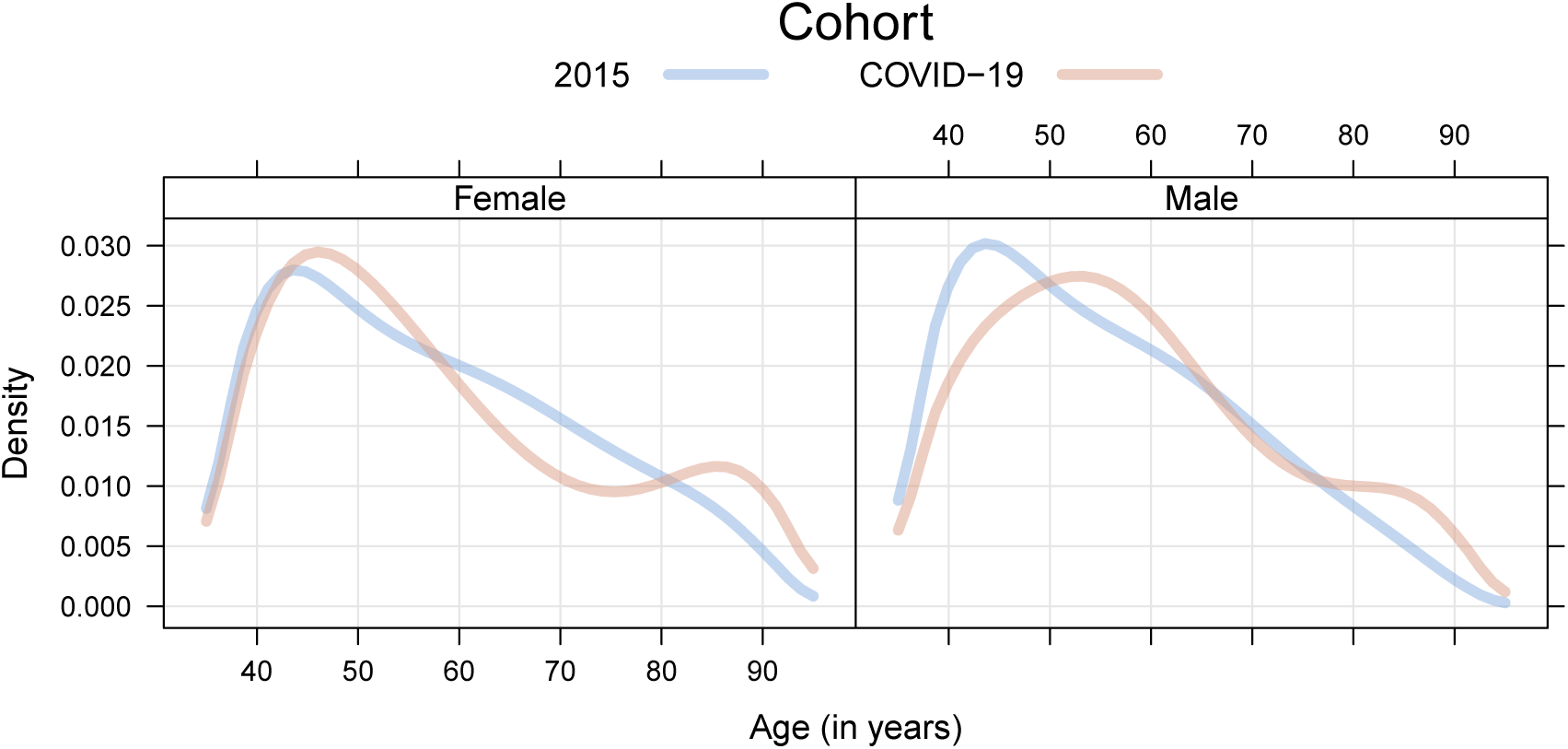
Swiss 2015 population and covid-19 cohorts. Comparison of age densities between the Swiss 2015 population and covid-19 cohorts, separately for females and males.

**Supplementary Figure 8:**
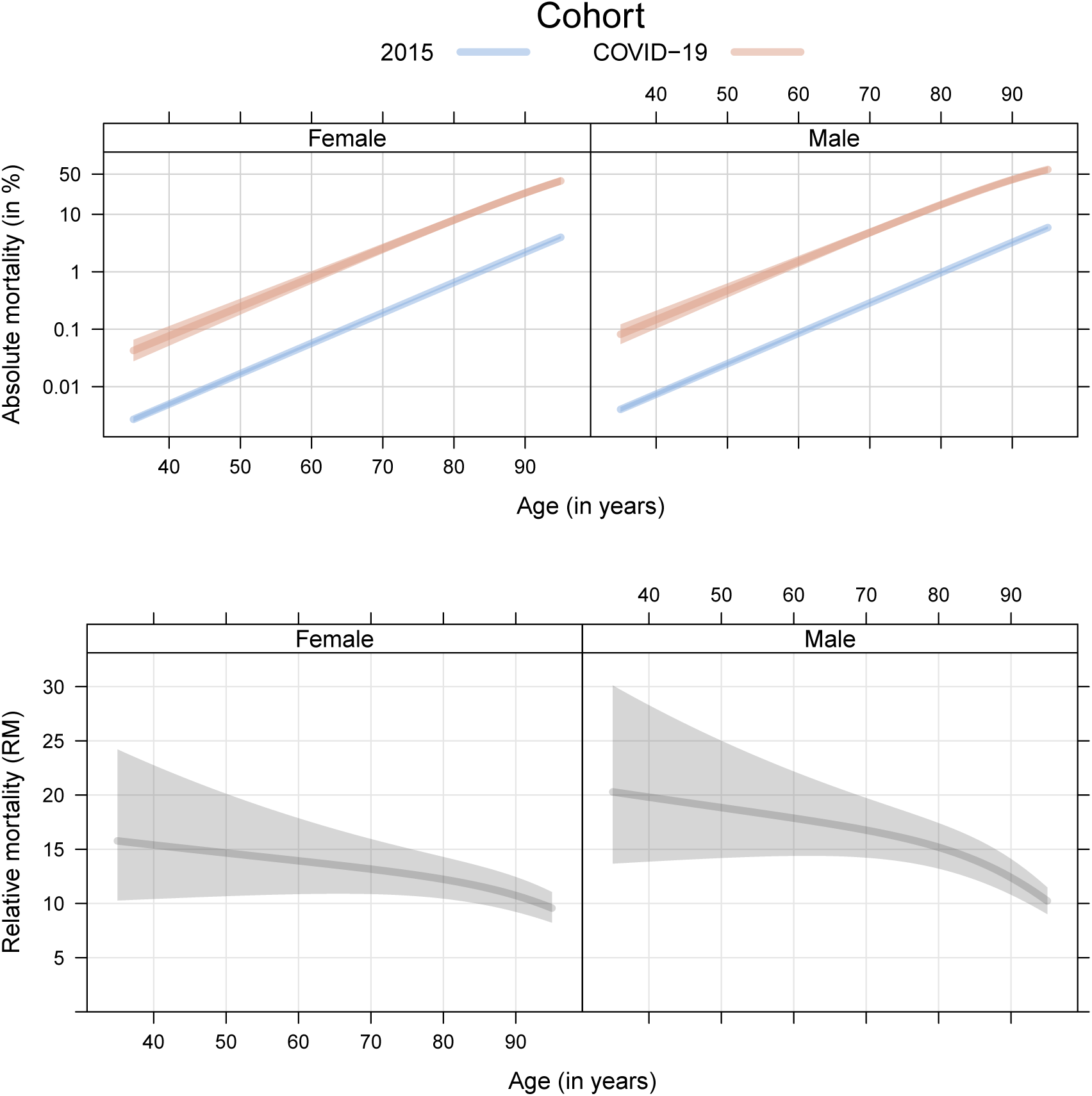
Swiss 2015 population and covid-19 cohorts. Comparison of absolute mortality (the probability of dying from any cause after 60 days, plotted on logarithmic scale) between the Swiss 2015 population and covid-19 cohorts, separately for females and males of different ages (top). Comparison of relative mortality (RM, the ratio of absolute mortalities in the covid-19 versus the 2015 cohort) between the two cohorts, for females and male of different ages (bottom). All estimates are plotted with 95% confidence bands.

**Supplementary Figure 9:**
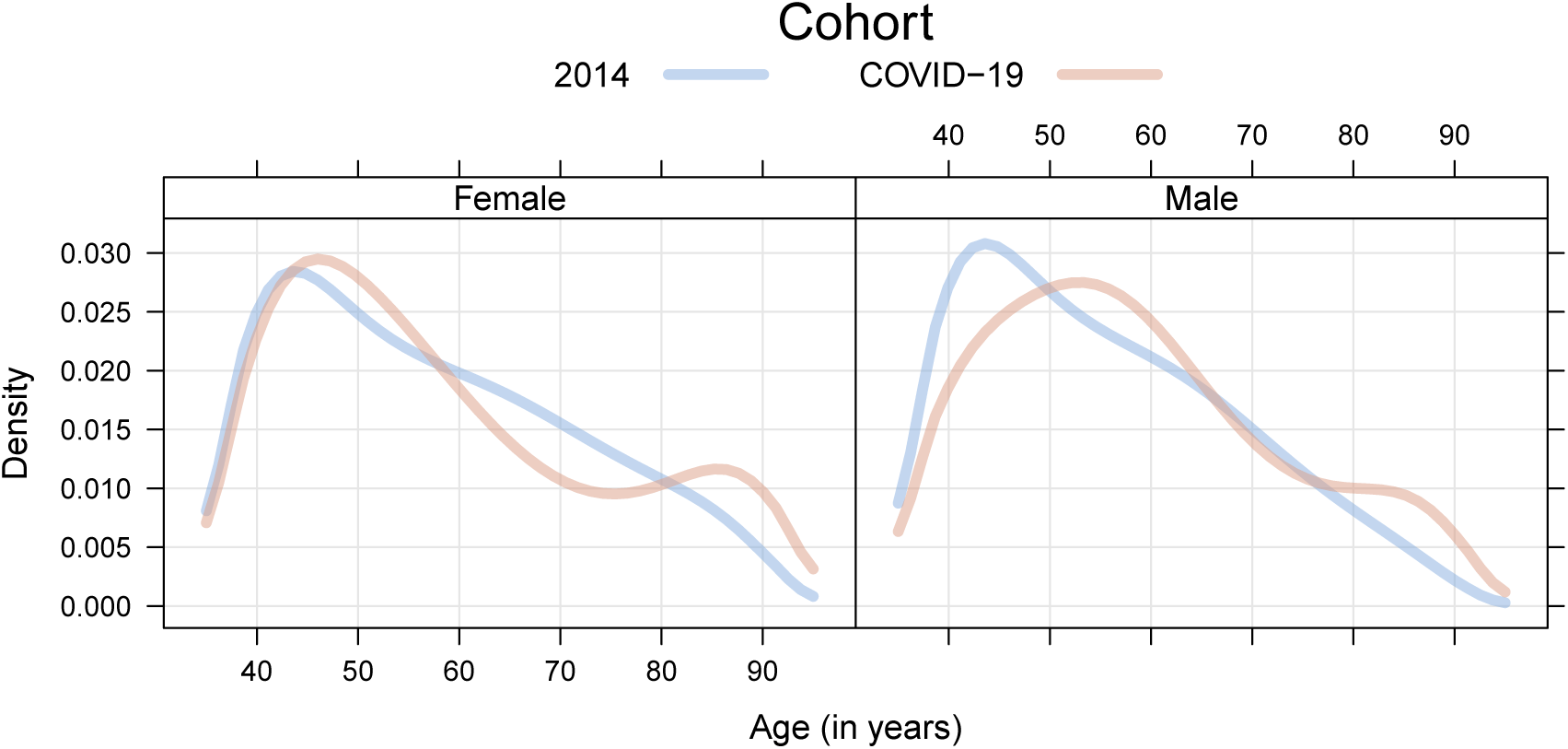
Swiss 2014 population and covid-19 cohorts. Comparison of age densities between the Swiss 2014 population and covid-19 cohorts, separately for females and males.

**Supplementary Figure 10:**
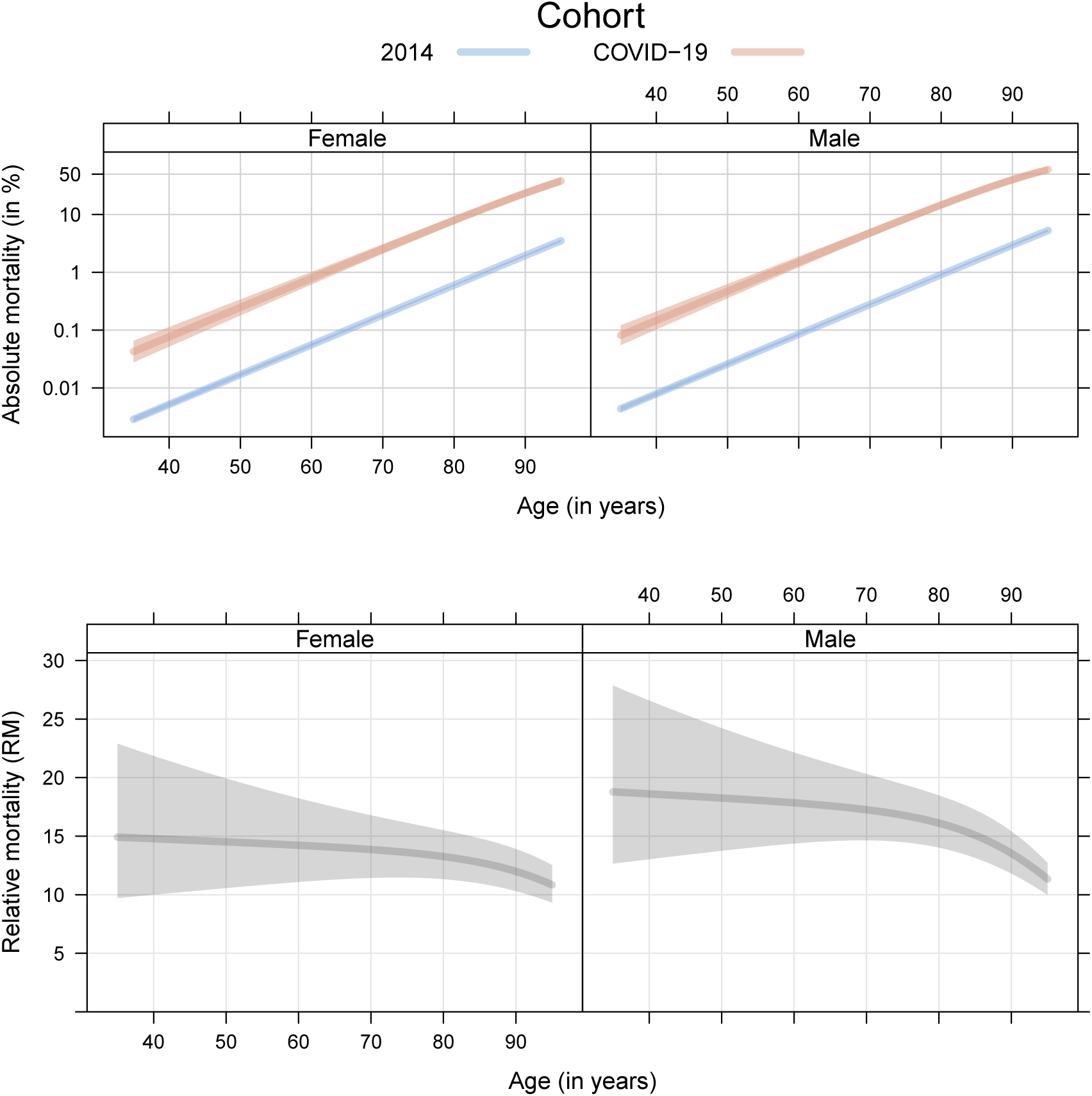
Swiss 2014 population and covid-19 cohorts. Comparison of absolute mortality (the probability of dying from any cause after 60 days, plotted on logarithmic scale) between the Swiss 2014 population and covid-19 cohorts, separately for females and males of different ages (top). Comparison of relative mortality (RM, the ratio of absolute mortalities in the covid-19 versus the 2014 cohort) between the two cohorts, for females and male of different ages (bottom). All estimates are plotted with 95% confidence bands.

## Footnotes

1 http://CRAN.R-project.org/package=mlt

2 http://CRAN.R-project.org/package=multcomp

## Notes

### Funding Statement

Swiss National Science Foundation. The funder of this study had no role in study design, data collection, data analysis, data interpretation, or writing of the final report.

### Summary of Updates

Unfortunately, some of the results reported in the initial version were incorrect due a coding error in the data preprocessing step. Despite our efforts to ensure computational validity by analysing the data independently in R and Stata, we failed to control all parts of the analysis. The relevant data preprocessing step merging population and patient data was only performed in R by the first author. This code assumed individual patient level data when in fact the information about population deaths had already been aggregated on a daily basis. Thus, only one death for a certain combination of sex and age was counted for a specific day, but multiple deaths were ignored. This mostly affected older ages, so the absolute population mortality for people older than 60 was underestimated. Consequently, the relative mortalities in these age groups were overestimated. The following conclusions were not affected by this correction: (1) Hazard increases linearly with age, both in the population and in patients. (2) The additional sex effect in patients seems is small and highly variable. (3) Relative mortality is at least as large as nine for all patients. (4) Age-based risk stratification deserves additional discussion in light of the absolute and relative results. The main conclusions remain intact and results in need to correction have been updated.

